# Adverse and benevolent childhood experiences and depression among women in rural Pakistan

**DOI:** 10.64898/2026.07.22.26358719

**Authors:** John A. Gallis, Yulu Pan, Rosalind J. Wright, Aparna G. Kachoria, Wan-Chen Lin, Allison Frost, Min Kyung Kim, Siham Sikander, Joanna Maselko

**Author notes:** **Corresponding author:** Min Kyung Kim.

## Abstract

Adverse childhood experiences (ACEs) are associated with maternal depression, including during the perinatal and postpartum periods when women face elevated risk. Benevolent childhood experiences (BCEs) may buffer negative effects of ACEs and promote mental health, yet evidence on their joint effects remains limited, particularly in low-resource settings. We investigated the joint effects of ACEs and BCEs on maternal depression from the third trimester of pregnancy through eight years postpartum among 804 women in rural Pakistan. Childhood experiences were characterized using both aggregated scores and latent class analysis to capture overall burden and item-level patterns. We tested the linear interaction term of ACEs and BCEs and utilized subgroup analysis to assess non-linear interactions. Overall, more than half (58.5%) of women experienced at least one ACE, and 6.2% of women experienced four or more ACEs. Commonly reported ACE domains were home violence (39.3%), neglect (19.7%) and family psychological distress (15.2%). Nearly half (45.3%) of women experienced all ten BCEs, and over half (51.5%) experienced 6-9 BCEs. Some BCE items had a very high prevalence, including liking themselves/feeling comfortable with themselves (96.6%), having at least one caregiver with whom they felt safe (96.5%), and having good neighbors (94.9%). Adjusting for ACEs, higher levels of BCEs were independently associated with fewer depressive symptoms (β = -0.25; 95% CI: -0.45, -0.05) and the promotive effects were consistent across ACE domains. Evidence of interaction suggested that BCEs buffered the adverse effects of ACEs among women with 1-3 ACEs (β = -0.56; 95% CI: -0.86, -0.26), but not among those with four or more ACEs. The associations between exposure to ACEs and depressive symptoms were weaker among women with higher levels of BCEs. Latent class analysis identified four distinct childhood experience profiles: Low-ACE/High-BCE (58.6%), High ACE/High BCE (19.5%), Low ACE/Low BCE (13.8%), High ACE/Low BCE (8.1%), with women in the High ACE/Low BCE class experiencing the greatest depressive symptom burden. Findings suggest that both adverse and benevolent childhood experiences are important for understanding maternal depression in low-resource settings, and informing interventions that reduce adversity exposure and foster positive developmental resources.

## Introduction

Maternal depression refers to a spectrum of depressive symptoms—such as persistent feelings of sadness or hopelessness—that can begin during pregnancy and persist for years, extending far beyond the typical perinatal period (i.e. pregnancy and the first 12 months postpartum) [1,2]. Depressive symptoms negatively affect mothers’ overall health, mother-child interaction, and child development [3]. The prevalence of maternal depression ranges from 17% to 30% across systematic reviews of global studies, with higher rates in low- and middle-income countries (LMICs) and low-resource rural settings [4–7]. A substantial proportion of women experience depressive symptoms for years after giving birth [1], which is further associated with mothers’ employment instability and economic hardship [8]. Consistent risk factors of maternal depression include limited social support, history of mental illness, adverse life events, and perceived stress [9,10].

Early-life experiences have also been linked with maternal mental health [11]. Adverse childhood experiences (ACEs) refer to negative psychosocial events occurring before 18 years old, including abuse, neglect, violence, and household dysfunction [12]. Greater exposure to ACEs is associated with increased risk of mood disorders, including depressive symptoms, post-traumatic stress, anxiety, and suicidal ideation in adulthood. [12–14]. According to a recent meta-analysis of 208 sample estimates from 22 countries, ACEs are very common, with around 60% of individuals having at least one ACE and 16% having at least four ACEs [15]. Benevolent childhood experiences (BCEs), as the counterpart to ACEs, capture nurturing relationships and supportive environments occurring before 18 years old [16]. In the original study that developed the BCEs scale among high-risk perinatal women, greater BCE exposure was associated with fewer traumatic stress disorder symptoms and stressful life events during pregnancy, independent of ACEs [16].

Although the adverse effects of ACEs are well established, ACEs do not deterministically lead to poor adult outcomes. Resilience theory emphasizes the role of BCEs such as positive external resources and individual assets that promote healthy development even in the context of childhood adversity [17,18]. For example, warm familial and extrafamilial support, and nurturing interpersonal relationships are linked to adaptive traits and build resilience, which help individuals overcome adversity later in life [19,20]. These promotive factors may also buffer the negative impact from earlier life risk factors [21].

ACEs and BCEs, as distinct and co-occurring dimensions of early childhood experiences, jointly shape long-term health outcomes and well-being [21,22]. Systematic reviews show a consistent promotive effect of BCEs on adult mental health outcomes even after accounting for ACE exposure [21,23,24]. In terms of the interactive effects of ACEs and BCEs, findings from studies that directly tested statistical interactions between ACEs and BCEs or used subgroup analyses are mixed. Some studies find no statistical interactive effect, and others suggest that the effect of BCEs varies across ACE levels and vice versa, depending on the population or setting [19,21,25].

Importantly, childhood experiences may differ not only in overall burden but also in domain-related content, with specific domains exerting distinct influences on mental health. For example, interpersonal violence within family and community was shown to be more strongly associated with depression than other forms of adversity [26,27]. Most existing studies operationalize ACEs and BCEs using aggregated total scores and model them as independent constructs in regression analyses. This approach implicitly assumes that every childhood experience produces the same impact on outcomes, and obscures heterogeneity across specific types and combinations of childhood experience [14]. Latent class analysis (LCA) offers a data-driven, person-centered approach that captures the patterns of co-occurring childhood experiences by identifying subgroups of individuals with similar profiles of ACEs and BCEs. However, very few studies have applied the LCA approach to examine ACEs and BCEs simultaneously [12,14,28,29]. In a study of 547 parents of 5 to 18-year-old children from the U.K., U.S., Canada, and Australia, Johnson et al. identified four latent classes: low-ACEs/high-BCEs, moderate-ACEs/high-BCEs, moderate-ACEs/low-BCEs, and high-ACEs/moderate-BCEs [14]. They found that moderate-to-high levels of ACEs were associated with elevated risk of mental health problems and family dysfunction, regardless of BCE level. Similarly, in an online survey sample of 488 adults in the U.K., Cain et al. identified four comparable classes (moderate ACEs/high BCEs, high ACEs/moderate BCEs, low ACEs/high BCEs, low ACEs/moderate BCEs), and found that individuals in the High ACEs/Moderate BCEs class experienced the worst psychological distress and suicidal thoughts and behaviors among all the classes [12].

Findings from these few studies highlight the need for further replication and extension using both traditional aggregated approaches and person-centered methods such as LCA to characterize childhood experiences. Furthermore, the majority of ACEs and BCEs studies have been conducted in Western and high-income settings, pointing to the need to assess generalizability to LMICs where childhood environment, resources, and stressors differ. In Pakistan, a LMIC in South Asia, the pooled prevalence of perinatal depression is estimated to exceed 30% [30,31], and most Pakistani women live in rural areas with low educational resources and face economic stressors. Prior cross-sectional studies from our study team linked ACEs with mental health outcomes at 36 months postpartum among women in rural Pakistan, particularly for ACE domains related to home violence and community violence, and family psychological distress [11,26]. However, it is unknown whether BCEs have a protective or promotive effect against the overall ACEs burden or specific domains of ACEs in this context.

In this study of 804 women in rural Pakistan, we aim to estimate the relationship between childhood experiences and maternal depression from the third trimester of pregnancy to eight years postpartum using both aggregated and LCA approaches. By examining effects of ACEs and BCEs simultaneously, we present how adverse and benevolent experiences jointly shape maternal depressive symptoms and identify populations with higher depression risks.

## Materials and Methods

### Study Population

This is a longitudinal analysis of 804 women from the Bachpan cohort study in rural Pakistan. The Bachpan study is a cluster randomized control trial designed to evaluate the impact of an adaptation of the Thinking Healthy Program, a peer-delivered community-based psychosocial intervention on maternal depression [32]. From October 2014 to February 2016, 1,154 pregnant women in their third trimester living in 40 village clusters were recruited and screened for depression using the Patient Health Questionnaire-9 (PHQ-9). Women who screened positive for depression (i.e., PHQ-9 score ≥10) were eligible to participate in the trial (N=570), and one of every three women who screened negative were enrolled as non-depressed reference group (N=584). Since baseline screening, participants were assessed with PHQ-9 at nine time points: baseline (third trimester during pregnancy), 3, 6, 24 months, and 3, 4, 6, 7, 8 years postpartum. Adverse Childhood Experiences (ACEs) were assessed at 3 years and Benevolent Childhood Experiences (BCEs) were assessed at the 7-year visit. This analysis used depressive symptoms measured by PHQ-9 from baseline to 8-year follow-up (nine timepoints) as the outcome measure, and ACEs/BCEs as exposure measures. Of the 1,154 women enrolled at the baseline screening, 265 missed the 3-year follow-up visit and 889 completed ACEs measures; 313 missed the 7-year follow-up visit and 841 completed BCEs measures.

### Ethics Statement

All participants were provided with a written consent form and voluntarily provided written informed consent. The Bachpan study received ethical approval from the institutional review boards at the University of North Carolina at Chapel Hill, Duke University, the Human Development Research Foundation, and the National Bioethics Committee, Pakistan.

### Exposures

Adverse childhood experiences (ACEs) were measured at 3 years postpartum using the Adverse Childhood Experiences - International Questionnaire ACE-IQ, a reliable instrument that has been adapted and validated across various social contexts [22,33]. The ACE-IQ was translated into Urdu and adapted for this study by removing the item on sexual abuse due to concerns of underreporting and potential risks to women after disclosing sexual abuse histories. Women were asked whether they had the following adverse experiences prior to age 18: home violence (physical abuse; emotional abuse; household member treated violently), family psychological distress (household member with alcohol/drug abuse; incarcerated household member; someone chronically depressed or mentally ill; parental death, parental separation or divorce), neglect (emotional neglect; physical neglect), and community violence (bullying; community violence; collective violence) (Table S1). For each item, response options included “yes” or “no” or “do not remember”. We coded “do not remember” (<2% for all the twelve individual items) as “no” so each ACE item was a binary indicator. For each of the four ACE domains, if a woman had at least one ACE item answering “yes”, the domain indicator was coded as “yes”. The total ACE score was calculated by summing the number of items that participant responded “yes”, and the score ranged from 0 to 12. The ACEs total score was also categorized into five groups: 0, 1, 2, 3, or ≥4 ACEs.

Benevolent childhood experiences (BCEs) were measured at 7 years postpartum using a 10-item benevolent childhood experiences scale, which was developed to assess positive early life experiences among adults with childhood adversities and validated in a sample of ethnically diverse, low-income pregnant women [16]. Women were asked whether they had the following positive experiences prior to age 18: (1) At least one caregiver with whom they felt safe; (2) At least one good friend; (3) Beliefs that gave comfort; (4) Enjoyment at school (5) At least one teacher that cared; (6) Good neighbors; (7) An adult (not parent or caregiver or the person from question 1) who could provide support or advice; (8) Opportunities to have a good time; (9) Like or feel comfortable with themselves; and (10) Predictable home routine, like regular meals and bedtime. For each item, response options included “yes” or “no” or “don’t know”. We coded “don’t know” (<2.5% for all the ten individual items) as “no” so each BCE item was a binary indicator. The total BCE score was calculated by summing the number of items that participant responded “yes”, and the score ranged from 0 to 10.

### Outcomes

The outcome of interest was maternal depressive symptom severity, measured using the Urdu version of Patient Health Questionnaire-9 (PHQ-9), a widely used screening tool with demonstrated acceptable validity and reliability among this population of pregnant women in Pakistan [34]. The PHQ-9 included nine items related to depressive symptoms and responses for each item included: 0 = not at all; 1 = several days; 2 = more than half the days; and 3 = nearly every day. The total score ranged from 0 to 27, with higher scores indicating greater depressive symptoms severity. Participants had assessment of PHQ-9 at nine time points: baseline (third trimester during pregnancy), 3, 6, 24 months and 3, 4, 6, 7, 8 years postpartum.

### Confounders

Confounders were determined a priori using a Directed Acyclic Graph based on previous knowledge (Figure S1). Confounders included maternal age (mean 26.6 years), immediate family’s history of mental illness (16.8%), and primary education completion (65.4%), all measured during the third trimester of pregnancy. Maternal age was treated as a continuous variable and family history of mental illness was a binary variable indicating whether any immediate family member had a mental illness. Primary education completion was coded as a binary variable indicating whether the women had passed more than 5 grades of education, and was used as a proxy for childhood socioeconomic status, as done previously [26].

## Statistical analysis

For descriptive analysis, we present the prevalence of individual ACE and BCE items, as well as the distribution of ACE and BCE total scores in the study population. Then, we used latent class analysis (LCA), a data-driven, person-centered approach to identify distinct latent classes based on the co-occurring patterns of childhood experiences using 12 ACE and 10 BCE binary items [14]. Polytomous variable latent class analysis was conducted using ‘poLCA’ function in ‘poLCA’ R package [35]. We compared models with two to five classes and selected the best-fitting model based on Bayesian Information Criterion (BIC) and entropy. Entropy is a standardized measure of classification certainty (range from 0 to 1) which was derived from individual posterior probabilities of class assignment, with higher values indicating better classification. We also checked class sample size to make sure there were no extremely small classes.

Because depressed women were oversampled during recruitment, we upweighted non-depressed women so that the study population could represent the underlying target population. Depressed women were assigned a sampling weight of 1. Non-depressed women in each cluster were assigned the same sampling weight, calculated as the inverse probability of enrollment among non-depressed pregnant women screened in that cluster, as done previously [26].

To account for informative missingness of ACEs and BCEs, we used stabilized inverse probability of censoring weights (IPCW) to upweight individuals in the study sample who share similar characteristics with those who were excluded [36]. Stabilized IPCW were calculated as Pr(C=0)/Pr(C=0|W), where C=0 indicates inclusion in the study sample and W included baseline confounders (maternal age, natal family’s history of mental illness and primary education completion) and baseline variables associated with censoring (with the p-value threshold of 0.15), including PHQ-9 score, trial arm, number of living children (first pregnancy, 1-3, 4 or more), family structure (nuclear family vs. joint or multiple household), current major depressive episode, household crowding (number of people per room), maternal disability measured by WHODAS score [37].

To examine the associations between ACEs, BCEs and maternal depressive symptoms, we fit linear-mixed effects models using PHQ-9 scores from nine timepoints as the outcome measure and different operationalizations of ACEs/BCEs as the exposure measure. Rather than averaging PHQ-9 scores across nine timepoints, we modeled repeated measures simultaneously in linear mixed-effects models. Time was included as a categorical fixed effect (nine timepoints) to model the non-linear changes of depressive symptoms. To account for correlation due to the clustered longitudinal design, we specified random intercepts at the village (cluster) level, and random intercepts and slopes for continuous timepoint at the individual level, with individuals nested within villages. Specifically, we first estimated the independent effects with only ACE total score or BCE total score included in the model (Model 0). As comparison, we estimated the independent effects of ACEs and BCEs using the ACE total score and BCE total score, adjusting for each other (Model 1). Then we added an interaction term of ACE and BCE total scores (Model 2). Given that the interaction may not be linear, we assessed BCE effects separately among women who had 0, 1-3, or ≥4 ACEs using three separate models (Model 3) and assessed ACE effects separately among women who had 10, 8-9, or ≤ 7 BCEs using separate models (Model 4). We further examined associations using latent classes derived from LCA as the exposure, with the low ACE/ high BCE class as the reference group (Model 5). Finally, we fit four separate models including BCE total score and one ACE domain indicator at a time (Model 6). Sampling weights and IPCW were multiplied together to generate the final weights, and robust (empirical) standard errors were used to account for clustering and weights. All analyses were performed using SAS 9.4 (SAS Institute Inc., Cary, NC) and R 4.4.2 (R Foundation for Statistical Computing, Vienna, Austria).

## Results

### Descriptive statistics

The study sample included 804 women who had complete measures for ACEs and BCEs. Baseline characteristics of 804 study participants and 350 excluded participants included in IPCW modeling were presented in Table S2. After applying sampling weights and IPCW, the weighted maternal age during pregnancy was 26.6 years, 68.5% had completed primary education, and 15.5% had an immediate family member with mental illness (Table S3).

The mean ACE total score was 1.2 (SD=1.4). Overall, 58.5% of women experienced at least one ACE, and 6.2% of women experienced four or more ACEs. Regarding ACE domains, 39.3% women experienced home violence, followed by neglect (19.7%), family psychological distress (15.2%), and community violence (7.2%). The mean BCE total score was 8.9 (SD=1.5). Nearly half of the women (45.3%) experienced all ten BCEs and over half (51.5%) experienced 6-9 BCEs, whereas 3.2% of women experienced five or fewer BCEs. Of the 10 BCE items, five had a prevalence of over 90%, including having at least one caregiver with whom they felt safe (96.5%), having beliefs that gave them comfort (91.5%), having good neighbors (94.9%), having opportunities to have a good time (92.8%), and liking themselves/ feeling comfortable with themselves (96.6%). The two least prevalent items—having at least one teacher that cared (79.1%) and having an adult (not parent or caregiver) who could provide support or advice (77.9%) were still reported by approximately 80% of women. The proportion of having experienced each ACE and BCE item is presented in Table 1 & Table 2.

**Table 1.**
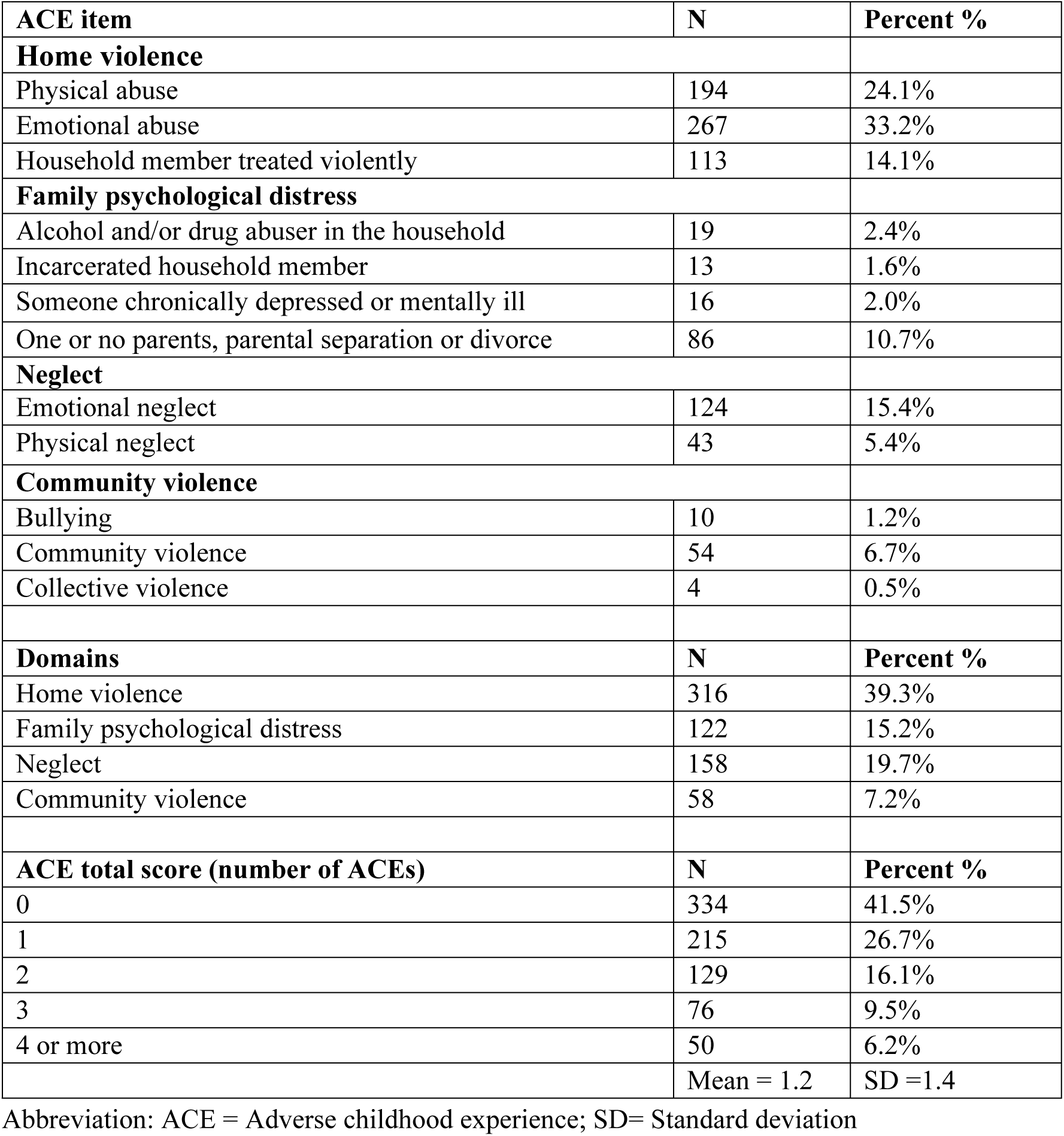
Adverse childhood experiences distributions.

**Table 2.**
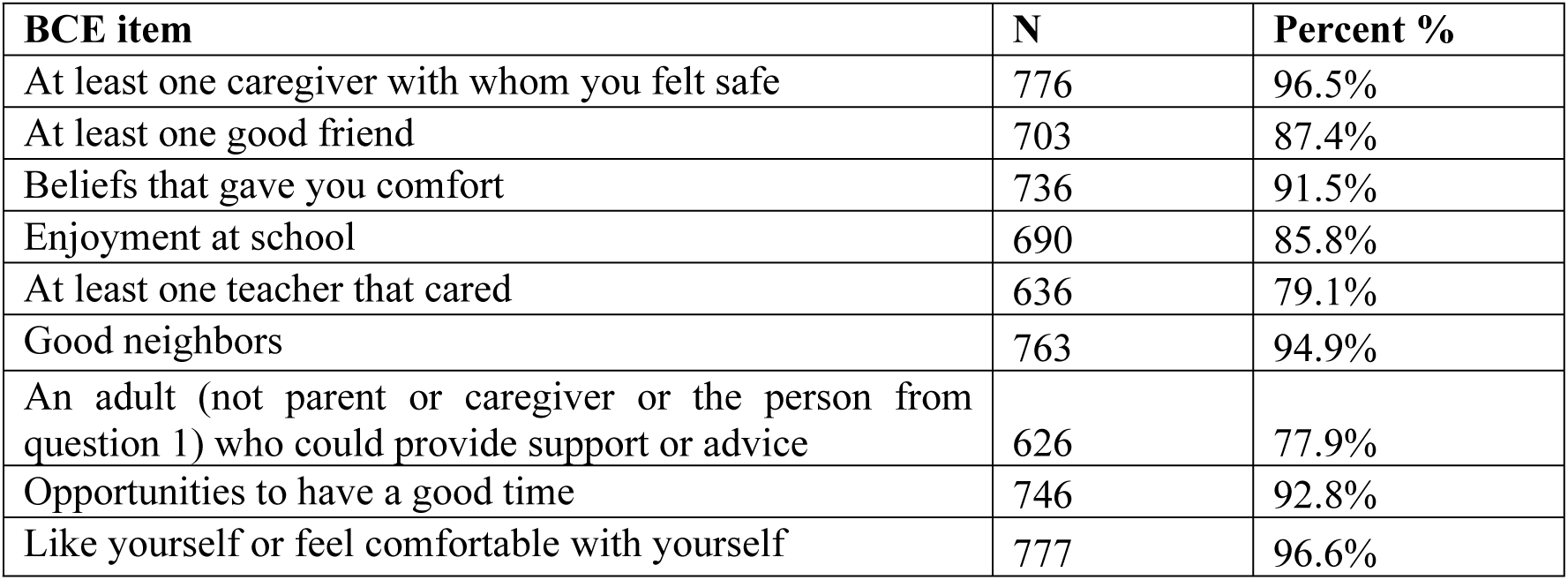
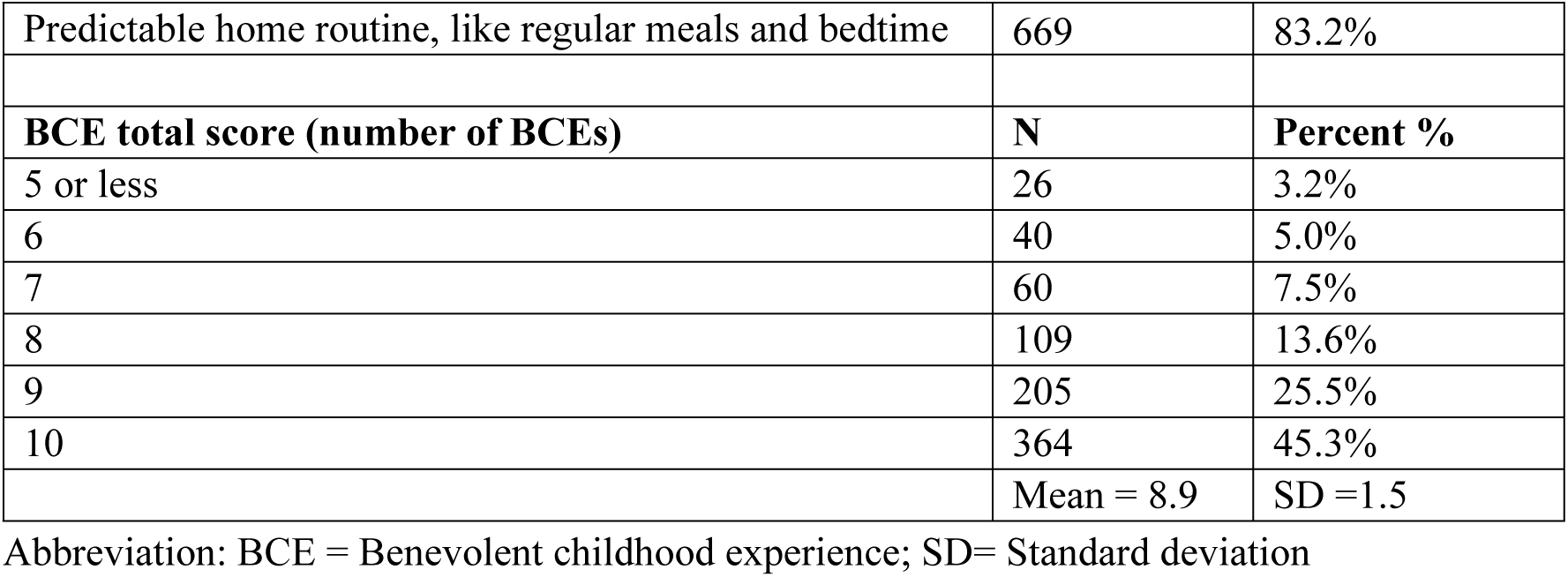
Benevolent childhood experiences distributions.

| BCE item | N | Percent % |
| --- | --- | --- |
| At least one caregiver with whom you felt safe | 776 | 96.5% |
| At least one good friend | 703 | 87.4% |
| Beliefs that gave you comfort | 736 | 91.5% |
| Enjoyment at school | 690 | 85.8% |
| At least one teacher that cared | 636 | 79.1% |
| Good neighbors | 763 | 94.9% |
| An adult (not parent or caregiver or the person from question 1) who could provide support or advice | 626 | 77.9% |
| Opportunities to have a good time | 746 | 92.8% |
| Like yourself or feel comfortable with yourself | 777 | 96.6% |
| Predictable home routine, like regular meals and bedtime | 669 | 83.2% |
| **BCE total score (number of BCEs)** | **N** | **Percent %** |
| 5 or less | 26 | 3.2% |
| 6 | 40 | 5.0% |
| 7 | 60 | 7.5% |
| 8 | 109 | 13.6% |
| 9 | 205 | 25.5% |
| 10 | 364 | 45.3% |
|  | Mean = 8.9 | SD = 1.5 |

Abbreviation: BCE = Benevolent childhood experience; SD= Standard deviation

### Latent class analysis

We compared the fitting metrics of latent class models with two to five classes (Table S4). The four-class model had the lowest BIC with an entropy of 0.78, reasonable class sizes, and meaningful classification based on averaged probability for ACE and BCE items (Figure 1 & Figure 2). Based on distinct patterns, we assigned descriptive labels to each class (Table S5). “High-ACE/ High-BCE” (n=157, 19.5%) was characterized by relatively high levels of both adverse and benevolent childhood experiences. “Low-ACE/Low-BCE” (n=111, 13.8%), was characterized by relatively low levels of both adverse and benevolent childhood experiences. “High-ACE/ Low-BCE” (n=65, 8.1%) was the least common one, with relatively high level of adverse childhood experiences and low level of benevolent childhood experiences. “Low-ACE/High-BCE” (n=471, 58.6%) was the most common class, with low level of adverse childhood experiences and high level of benevolent childhood experiences.

**Figure 1.**
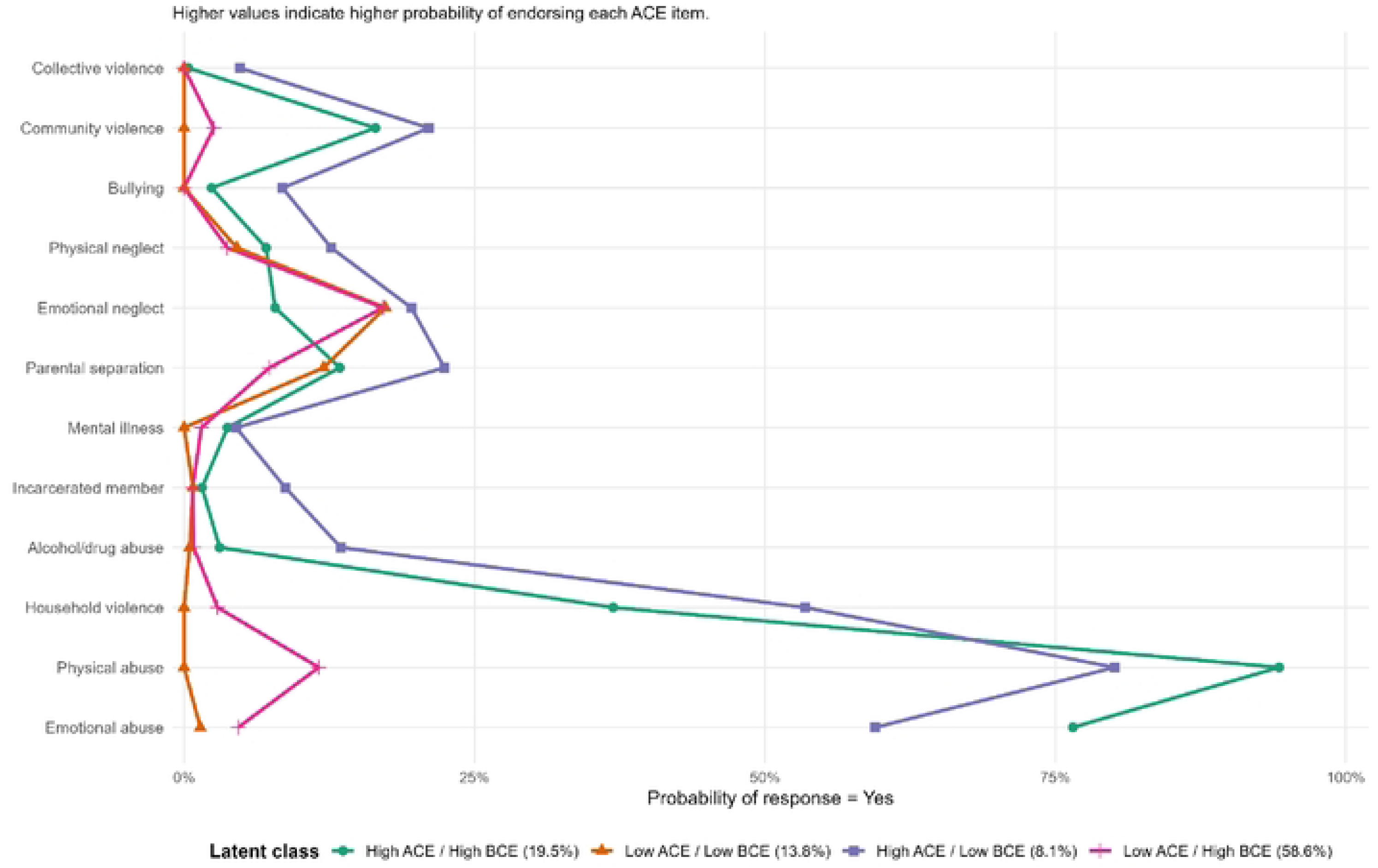
Probability of Reporting ACE items by latent class. Y axis indicates the probability of reporting each of the 12 ACE items by four latent class groups, including High-ACE/High-BCE (n=157, 19.5%), Low-ACE/Low-BCE” (n=111, 13.8%), High-ACE/ Low-BCE” (n=65, 8.1%), Low-ACE/High-BCE” (n=471, 58.6%).

**Figure 2.**
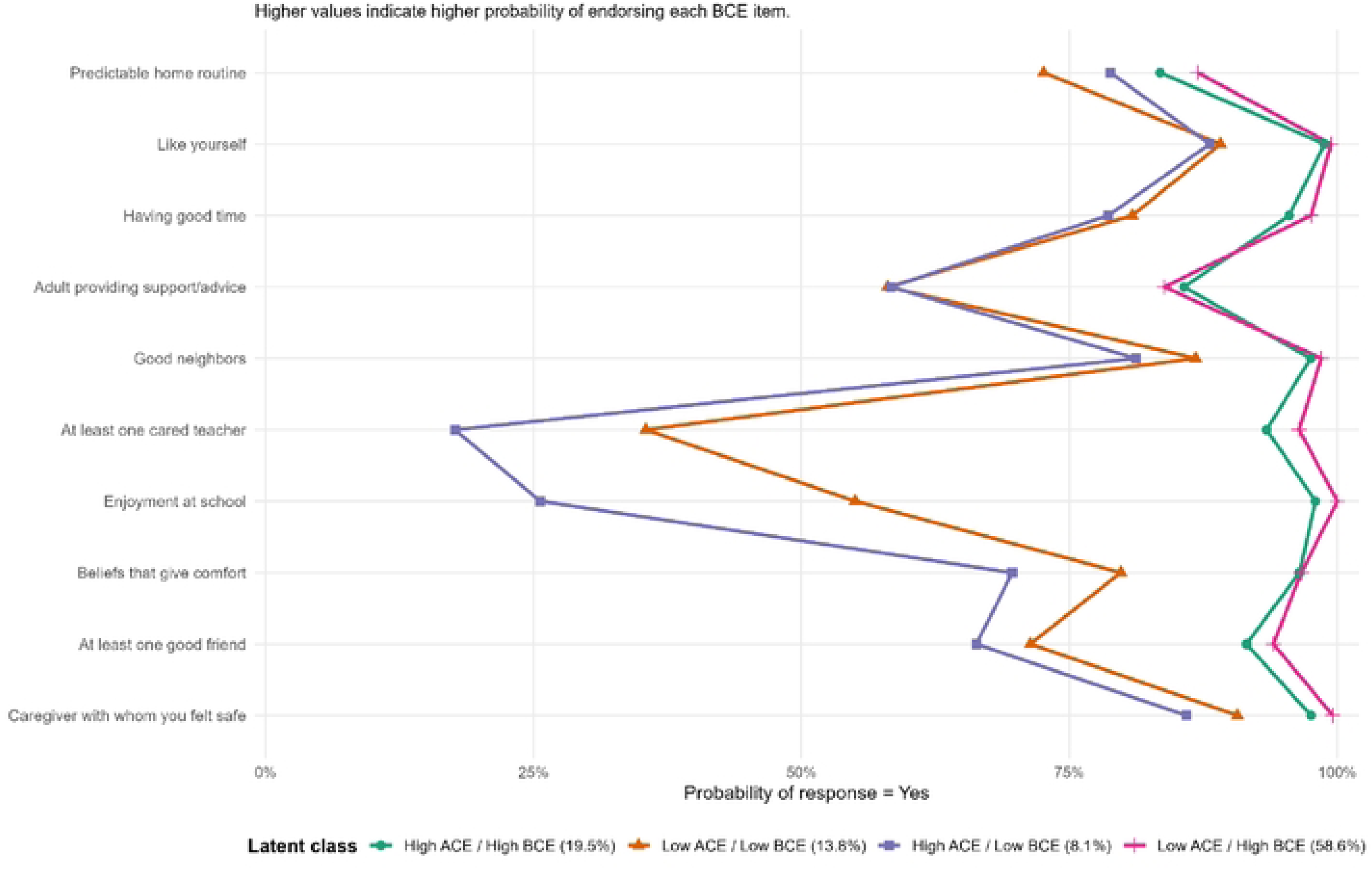
Probability of Reporting BCE items by latent class. Y axis indicates the probability of reporting each of the 10 BCE items by four latent class groups, including High-ACE/High-BCE (n=157, 19.5%), Low-ACE/Low-BCE” (n=111, 13.8%), High-ACE/ Low-BCE” (n=65, 8.1%), Low-ACE/High-BCE” (n=471, 58.6%).

Class separation was largely driven by a subset of ACE and BCE items that clearly distinguished high versus low exposure patterns. Women classified into the High-ACE classes (i.e. High-ACE/High-BCE and High-ACE/Low-BCE) exhibited substantially higher probabilities of exposure to physical abuse, emotional abuse, household violence, and community violence compared with women in the Low-ACE classes, while prevalence differences for other ACE items were relatively small (Fig 1A). In contrast, women classified into the Low-BCE classes (i.e. High-ACE/Low-BCE and Low-ACE/Low-BCE) consistently reported lower probabilities across all BCE items relative to High-BCE classes, with particularly pronounced differences for enjoyment at school and having at least one caring teacher (Fig 1B). These item-level patterns suggest that certain highly discriminating ACE and BCE items may play a central role in defining the latent classes.

### Childhood experiences and depressive symptom severity

Both adverse and benevolent childhood experiences were independently associated with maternal depressive symptoms (Table 3). In Model 1 adjusting for BCEs and confounders, each additional ACE exposure was associated with higher average PHQ-9 score across all follow-up waves (β =0.44, 95% CI: 0.24, 0.64). In contrast, adjusting for ACEs and confounders, each additional BCE exposure was associated with lower PHQ-9 average score across all follow-up waves (β = -0.25, 95% CI: -0.45, -0.05). The independent effects for ACEs and BCEs adjusting for each other in Model 1 were similar to those in Model 0 that only included ACEs (β = 0.46, 95% CI: 0.26, 0.66) or only included BCEs (β = -0.30, 95% CI: -0.50, -0.09). The ACE×BCE interaction term was not significant in Model 2, but subgroup analysis (Models 3-4) indicated that the protective effect of BCEs varied by the level of ACE exposure, and harmful effect of ACEs also varied by the level of BCE exposure. BCE exposure was associated with lower PHQ-9 scores among women with 1-3 ACEs (β = -0.56, 95% CI: -0.86, -0.26), while the protective effect of BCEs was close to none among women who had no exposure to ACEs (β = 0.02, 95% CI: -0.27, 0.30) or women who had ≥ 4 ACEs (β = -0.06, 95% CI: -0.84, 0.72). Exposure to ACEs was consistently associated with higher PHQ-9 scores regardless of BCEs exposure but had a stronger effect among women with lower level of BCE exposure. In Model 5 using latent classes as exposure, compared with women in Low-ACE/High-BCE reference group, women in High-ACE/High-BCE, Low-ACE/ Low-BCE, High-ACE/ Low-BCE had 0.92 (95% CI: 0.13, 1.70), 0.41 (95% CI: -0.46, 1.28), and 2.73 (95% CI: 1.61, 3.86) point higher average PHQ-9 scores, respectively. Regarding the effect of ACE domains (Model 6), home violence (β = 0.89, 95% CI: 0.25, 1.52), family psychological distress (β = 0.84, 95% CI: 0.03, 1.65), community violence (β = 0.90, 95% CI: -0.27, 2.07), and neglect (β = 0.46, 95% CI: -0.32, 1.24) were all associated with higher PHQ-9 scores even though some estimates were not statistically significant. Across all ACE domains, BCEs consistently showed a protective effect on depressive symptoms, with each additional BCE exposure associated with around 0.29 point lower PHQ-9 scores.

**Table 3.** Associations between ACE, BCE and depressive symptom severity (PHQ-9)

| Model | Predictor | Beta coefficient | 95% CI | p-value |
| --- | --- | --- | --- | --- |
| *Model 0: Include ACE or BCE only* |  |  |  |  |
| Include ACE only | **ACE total** | 0.46 | (0.26, 0.66) | <0.001 |
| Include BCE only | **BCE total** | -0.30 | (-0.50, -0.09) | 0.006 |
| **Model 1: Main effect** |  |  |  |  |
| Adjust for BCE | **ACE total** | 0.44 | (0.24, 0.64) | <0.001 |
| Adjust for ACE | **BCE total** | -0.25 | (-0.45, -0.05) | 0.01 |
| **Model 2: Add interaction term** |  |  |  |  |
| Include ACE, BCE, and interaction | **ACE total** | 1.09 | (-0.01, 2.18) | 0.05 |
|  | **BCE total** | -0.15 | (-0.39, 0.10) | 0.25 |
|  | **ACE \* BCE** | -0.08 | (-0.21, 0.05) | 0.25 |
| **Models 3: Subgroup analysis by ACE** |  |  |  |  |
| ACE = 0 | **BCE total** | 0.02 | (-0.27, 0.30) | 0.90 |
| ACE = 1-3 |  | -0.56 | (-0.86, -0.26) | <0.001 |
| ACE ≥ 4 |  | -0.06 | (-0.84, 0.72) | 0.88 |
| **Models 4: Subgroup analysis by BCE** |  |  |  |  |
| BCE = 10 | **ACE total** | 0.33 | (-0.01, 0.67) | 0.06 |
| BCE = 8-9 |  | 0.41 | (0.07, 0.75) | 0.02 |
| BCE ≤ 7 |  | 0.59 | (0.10, 1.07) | 0.02 |
| **Model 5: Latent class (Low-ACE/ High-BCE as reference)** |  |  |  |  |
| Latent class categories | **High ACE/High BCE** | 0.92 | (0.13, 1.70) | 0.02 |
|  | **Low ACE/Low BCE** | 0.41 | (-0.46, 1.28) | 0.35 |
|  | **High ACE/Low BCE** | 2.73 | (1.61, 3.86) | <0.001 |
| **Models 6: ACE domain and BCE total** |  |  |  |  |
| ACE = Home violence | **Home violence** | 0.89 | (0.25, 1.52) | 0.006 |
|  | **BCE total** | -0.28 | (-0.48, -0.08) | 0.006 |
| ACE = Family psychological distress | **Family psychological distress** | 0.84 | (0.03, 1.65) | 0.04 |
|  | **BCE total** | -0.28 | (-0.49, -0.07) | 0.01 |
| ACE = Neglect | **Neglect** | 0.46 | (-0.32, 1.24) | 0.25 |
|  | **BCE total** | -0.29 | (-0.50, -0.08) | 0.007 |
| ACE = Community violence | **Community violence** | 0.90 | (-0.27, 2.07) | 0.13 |
|  | **BCE total** | -0.29 | (-0.50, -0.09) | 0.006 |

All models used mixed effect linear models with weights (combination of sampling weight and IPCW) and accounted for clustering using robust standard errors. All models adjusted for maternal age, primary education completion, and mental illness in immediate family. All models included individual random effect for intercept and continuous time slope, and cluster-level random intercept. Time was included as a categorical fixed effect in the model.

## Discussion

To our knowledge, this is the first investigation of joint patterns of ACEs and BCEs in Pakistan. Using both aggregated total scores and person-centered latent class approaches, we examined associations between childhood experiences and maternal depressive symptoms from pregnancy to eight years postpartum. When modeled as aggregated scores, higher ACE exposure was independently associated with greater depressive symptoms, whereas higher BCE exposure was independently associated with fewer depressive symptoms. Subgroup analyses suggested interactive effects of ACE and BCE. Specifically, the protective effect of BCEs varied by the level of ACE exposure, with effects approaching the null among women with no ACEs or with four or more ACEs. Conversely, the harmful effect of ACEs showed a dose-response relationship, with stronger associations observed among women with lower levels of BCEs. Findings from latent class analysis further corroborated these patterns. Women in High-ACE/ Low-BCE experienced the greatest depressive symptom burden, followed by those in the High-ACE/High-BCE and Low-ACE/ Low-BCE classes, suggesting that the co-occurrence of high childhood adversity and limited benevolent experiences exerts the highest risk for maternal depression.

### Distribution pattern of ACEs and BCEs

Overall, childhood adversity was common among Pakistani women in this study, with 58.5% reporting at least one ACE. The prevalence was higher than estimates reported in most high-income countries, but lower than those in some other LMICs such as Kenya (65.8%), Malawi (almost 100%) and Mexico (almost 100%) [38–40]. The relatively lower prevalence observed in our sample may be due to the exclusion of sexual abuse items and underreporting of child abuse and family psychological distress in collectivist contexts, where such experiences may be stigmatized [41]. However, direct comparisons of ACE prevalence across countries should be interpreted cautiously because studies differ in measurement, sampled populations, and reporting context. The ACE prevalence varied substantially across domains and individual events, with home violence (including physical abuse, emotional abuse, and household member treated violently) emerging as the most common domain (39.3%).

In contrast, the BCE prevalence was high in our sample, with a mean of 8.9 items, similar to the levels reported in previous studies [25]. The distribution of individual BCE items has been observed to vary across populations and cultural contexts [16,25,42]. For example, the prevalence of having good neighbors and liking oneself during childhood was higher in our sample and in studies from China than in studies from Western countries [16,43–45]. The high prevalence of several BCE items in our cohort is encouraging, as it suggests that many women had access to social and interpersonal resources beyond their immediate family, including support from neighbors, friends, teachers, non-caregiver adults, and from their own beliefs and positive self-concept. These external and internal resources may support mental health by reducing social isolation, promoting adaptive coping strategies, and fostering a sense of belonging and purpose in the context of childhood adversity [16,46,47].

### Latent class analysis (LCA)

Using a person-centered LCA approach, we identified four latent classes that demonstrated the best model fit. This is consistent with previous studies applying LCA to individual ACE and BCE items [12,14], supporting the notion that ACEs and BCEs could co-occur and capture distinct dimensions of early life experiences. More than half of the sample was classified into Low-ACE/High-BCE (n=471; 58.6%), representing a nurturing developmental environment characterized by low exposure to childhood adversity alongside abundant positive experiences. In contrast, a small subset of women (n=65; 8.1%) fell into the High-ACE/ low-BCE class, representing the most vulnerable profile. This group experienced high levels of childhood adversity in the absence of compensatory benevolent resources and had the highest burden of depressive symptoms.

High-ACE/ High-BCE and Low-ACE/ Low-BCE classes also exhibited elevated depressive symptoms, but to a lesser extent than the High-ACE/ Low-BCE class. When comparing High-ACE classes versus Low-ACE classes, women in High-ACE classes did not have a high prevalence uniformly across all ACE items, but had a relatively high probability in specific items, particularly those related to home violence. In contrast, distinctions between Low-BCE classes and High-BCE classes were more pronounced and consistent across items, with consistently lower probability of reporting each item in the Low-BCE classes, especially for schooling-related experiences (i.e. at least one caring teacher and enjoyment at school). These results suggest that while childhood adversity may cluster within particular domains, benevolent experiences tend to reflect a broader set of resources and social networks that operate in combination to promote resilience [21].

### Independent effects of BCEs and ACEs

In terms of associations between childhood experiences and depressive symptoms, we found that adverse and benevolent childhood experiences had independent effects. In our previous work, higher ACE total score was associated with greater depressive symptoms cross-sectionally at three years postpartum [26]. We extended this work by demonstrating that the relationship persists when accounting for BCEs and when depressive symptoms were assessed longitudinally from pregnancy through eight years postpartum. After accounting for covariates and ACE exposure, a higher BCE total score remained associated with fewer depressive symptoms. Similar findings have been reported across diverse populations and contexts, indicating that higher BCEs independently predict better mental health even after controlling for ACEs [21].

The direct promotive effect of BCEs on depressive symptoms is consistent with the Compensatory Model of resilience theory—individuals’ resources and assets operate as compensatory factors that directly promote better mental health outcomes independent of risk exposure [17]. However, not all studies found the independent effects after controlling for one another, as in our study. In two studies of pregnant women in the US (mean age 30 and 29 years old), ACEs or BCEs no longer yield significant associations with maternal depressive symptoms during prenatal period after adjusting for covariates [48,49].

At the domain level, community violence, home violence, and family psychological distress exhibited stronger associations with depressive symptoms than neglect after accounting for BCEs. The deleterious mental health effects of violence and family distress have been consistently documented in prior literature [27,50,51]. ACE domains vary by context and may affect adult mental health through different biological, psychological, and social pathways [52]. Notably, the promotive effect of BCEs was consistent across four ACE domains, suggesting potential utility of interventions that provide positive resources against all types of ACEs.

### Interactive effects of BCEs and ACEs

To test whether the effect of BCEs on depressive symptoms varied by level of ACE exposure, we first fitted a model with ACE×BCE interaction term, which did not support a significant interactive effect. Prior studies have reported mixed findings regarding interactive effects. Some studies identified significant interaction terms for outcomes other than depression [53,54], while others reported null interactions for depressive symptoms [55]. In subgroup analyses stratified by BCE levels (10, 8-9, ≤7), ACE exposure remained significantly associated with depressive symptoms across all strata, but the associations were weaker among women with higher levels of BCEs. Another two studies found more encouraging results that the ACEs were associated with negative outcomes only among individuals with low to moderate BCE levels, and negative effects no longer exist when BCEs levels were high [56,57]. These patterns align with the Protective Factor Model of resilience theory, which posits that promotive factors such as BCEs modify the relationship between risk and outcomes [17,18]. BCEs may influence the mental health outcomes through coping mechanisms selection that mediate responses to adversity and stress. For example, people with higher BCEs are more likely to use adaptive coping strategies such as seeking social support and less likely to use maladaptive strategies such as self-blame and social withdrawal [47]. In our study, the observed dose-response attenuated relationship between ACEs and depressive symptoms as BCEs level increases indicates that BCEs buffer or neutralize the negative effects of ACEs. Some other studies did not support the Protective Factor Model as they found that ACEs had a stronger effect on worse outcomes among people with high BCEs levels [19,58].

We also conducted subgroup analysis to assess the effect of BCEs across ACEs levels (0, 1-3, or ≥4). Among women with 1-3 ACEs, higher BCE total score was associated with fewer depressive symptoms, whereas the association was close to null among people with no ACE or with four or more ACEs. In Bethell et al. study, the BCE association with depression remained stable across ACE exposure level [46], while most other studies found stronger BCE effects in the context of lower childhood adversity [19,45,58,59]. The null effect among people with no ACE exposure may reflect a ceiling effect, as these people generally had high BCE levels and little variations in depressive symptoms. The null effect among people with four or more ACEs would indicate another scenario when the ACEs are too harmful and overwhelms the protective benefits of BCEs. The Challenge Model of resilience theory suggests that exposure to modest levels of adversity may facilitate the development of coping mechanisms against future adverse exposures [17,18]. However, if the adversity is too taxing or overwhelming, it can hinder the building of resilience [17,18]. In our study, women reporting a high number of ACEs (e.g. ≥4) may have experienced such a severe level of adversities that even the existence of BCEs was insufficient to offset the harmful effect.

## Strength and Limitations

This study is the first to investigate the joint effects of ACEs and BCEs on maternal depression in Pakistan, South Asia. We employed both a traditional aggregated approach and a person-centered latent class analysis to characterize overall and item-specific patterns of childhood experiences. Additionally, we utilized subgroup analysis to assess non-linear interactions between ACEs and BCEs that may not be detected through conventional modeling of interaction term. These approaches provide insights into how the co-occurring patterns of ACEs and BCEs jointly shape maternal depressive symptoms.

Several limitations should be noted. First, ACEs and BCEs were reported retrospectively in adulthood, which may introduce recall bias. In addition, ACEs and BCEs were measured at follow-up visits rather than concurrently with depressive symptom assessments, which may affect temporal comparability between exposures and outcomes. Women’s mental health state may also affect how participants report ACEs and BCEs, leading to differential misclassification. For example, women experiencing depression may be more likely to recall or report adverse experiences. Second, we had limited data on childhood-specific covariates. We used women’s education attainment as a proxy for their childhood socioeconomic status. There may be other unmeasured confounders, such as parenting practices and familial expectations during women’s childhood. Third, we had loss to follow-up and missingness for ACEs and BCEs, but we used stabilized IPCW to account for informative censoring. Lastly, the ACE-IQ may not fully capture adversities during childhood in the Pakistan context, and the sexual abuse item was excluded due to aforementioned concerns. Given the prevalence of child marriage and sexual violence in Pakistan [60], the exclusion of sexual abuse may underestimate the prevalence of ACEs and the associations with maternal depression.

## Conclusions

While ACEs have been a well-established risk factor for poor mental health, our work demonstrates that BCEs exert both independent promotive effects and context-dependent protective (interactive) effects on maternal depressive symptoms. Our findings support multiple resilience theory frameworks, suggesting that BCEs operate as promotive factors that directly reduce depressive symptoms (Compensatory Model), and also serve as protective factors that buffer the negative effects of ACEs when adversity is not overwhelming (Protective Factor Model & Challenge Model). Our study highlights the importance of examining both ACEs and BCEs when studying childhood experiences. Programs addressing childhood experiences should prioritize both reducing exposure to adversity and fostering positive developmental resources. Positive experiences may be promoted through supportive family and peer relationships, community engagement opportunities, equitable school environments, and interventions that support emotional growth and adaptive coping skills. Additional psychosocial support is critical for children exposed to high levels of adversity, as for them benevolent experiences alone may be insufficient to offset risks.

## Data Availability

Data will be made available on request.

## Acknowledgements

The authors would like to thank the team at the Human Development Research Foundation (HDRF) and the National Bioethics Committee Pakistan, including Siham Sikander (Bachpan Project leader), Rakshanda Liaqat, Tayyiba Abbasi, Maria Sharif, Samina Bilal, Quratul-Ain, Anum Nisar, Amina Bibi, Shaffaq Zufiqar, Sonia Khan, Ahmed Zaidi, Ikhlaq Ahmad, and Najia Atif for their meaningful contributions to the study’s design and implementation. We also gratefully acknowledge the larger Bachpan and SHARE study teams. Lastly, we are deeply grateful to the contribution of women, children, and communities that are a part of the Bachpan cohort.

## Supporting information

**Figure S1. Directed acyclic graph of childhood experiences and maternal depressive symptoms**

**Table S1. ACE questionnaire items and domains**

**Table S2. Baseline Characteristics in IPCW model**

**Table S3. Weighted characteristics statistics after applying sampling weights and IPCW**

**Table S4. Latent class analysis of adverse and benevolent childhood experiences model fitting metrics**

**Table S5. Latent classes labels and distribution**

## Notes

### Competing Interest Statement

The authors have declared no competing interest.

